# Implementation and first results of the Czech nationwide prostate cancer screening pilot programme

**DOI:** 10.64898/2026.01.05.26343424

**Authors:** Marcela Koudelková, Kateřina Hejcmanová, Marek Babjuk, Roman Zachoval, Jiří Ferda, Ola Bratt, Renata Chloupková, Ondřej Ngo, Karel Hejduk, Vlastimil Válek, Ladislav Dušek, Ondřej Májek

## Abstract

**Background:** In 2022, the European Union recommended piloting organised prostate cancer screening. Subsequently, the Czech Republic launched a nationwide pilot programme fully reimbursed by public health insurance in January 2024 to reduce the high incidence of late-stage prostate cancer and the widespread unorganised prostate-specific antigen (PSA) testing.

**Objective:** To describe the strategy, methodology, and first results of the Czech nationwide prostate cancer screening pilot programme.

**Methods:** We analysed data from the first 12 months of the programme, which targets men aged 50–69 years without a previous prostate cancer diagnosis. General practitioners and urologists offer men an initial PSA test. Those with PSA ≥3.0 µg/l are referred to certified urologists, based on urology assessment selected for magnetic resonance imaging (MRI) and, if indicated, targeted biopsy. Reported key indicators are PSA testing coverage and overall use of pre-biopsy MRI.

**Key Findings and Limitations:** Screening was offered to 150,498 men; PSA results were available for 146,109 (97.1%), 8.8% of whom had ≥3.0 µg/l. Within 6 months, 67.2% with PSA ≥3.0 µg/l underwent urological evaluation. From 2023 to 2024, the 2-year PSA testing rate rose from 47.4% to 53.3%, and pre-biopsy MRI use from 28.0% to 38.3% (10.3 percentage-point difference, 95% CI 8.3–12.2). Data on diagnostic outcomes are not yet analysed.

**Conclusions and Clinical Implications:** These results demonstrate the feasibility of transforming widespread unorganised PSA testing to an organised screening programme with improved adherence to the recommended diagnostic pathway. Further evaluation to assess outcomes and efforts to reduce opportunistic testing will follow.

**Advancing Practice:** *What does this study add?:* This is the first population-level evaluation of a nationwide organised prostate cancer screening programme in Central Europe. The findings demonstrate the feasibility of integrating a structured screening programme with PSA testing in routine primary care into a national health system. The programme improved adherence to guideline-based diagnostic pathways.

*Patient Summary:* This report presents the initial results of a new nationwide, organised prostate cancer screening programme in the Czech Republic. The programme contributed to increased screening coverage in the target population (50-69 years) and improved the use of MRI before biopsy, which is now the recommended practice.

## Introduction

Prostate cancer is one of the most common malignancies among men in the European Union, and the incidence is steadily increasing [1]. Late diagnosis is a persistent challenge, with approximately one-third of cases detected in stage III or IV in the Czech Republic [2], when therapeutic options are limited and the prognosis is poor [2]. In contrast, early detection significantly increases the chance of curative treatment and long-term survival [3] [4]. Approximately 8,000 new prostate cancer cases are diagnosed annually in the Czech Republic, nearly three times more than two decades ago [2]. This rise is attributable not only to the ageing population but also to widespread use of opportunistic prostate-specific antigen (PSA) testing. International randomized trials have demonstrated that organised PSA-based screening can reduce prostate cancer mortality [4]. However, it also carries risks of overdiagnosis and overtreatment, although less so than unorganised (opportunistic) testing [4] [5]. Screening may thus lead to clinically unjustified diagnostic interventions and put an unnecessary burden on healthcare services [6]. In response to current challenges and the potential benefits of organised screening using modern prostate cancer diagnostics, including magnetic resonance imaging (MRI) and targeted biopsy, the EU in 2022 recommended that member states assess the feasibility and effectiveness of organised screening programmes [8]. This initiative was also endorsed by the 2022 International Prostaforum declaration, which supported the launch of pilot programmes for early detection across Europe [9].

Based on these international and national frameworks, a nationwide prostate cancer screening pilot programme was launched in the Czech Republic in January 2024 [10]. The primary objective of the programme is to assess the feasibility of organised prostate cancer screening using PSA as a simple and cost-effective biomarker to guide diagnostics and reduce advanced-stage disease. A further aim is to validate the structured early detection algorithm incorporating PSA-based risk stratification and MRI-guided biopsy within the Czech healthcare system [11].

We here describe the strategy and methods employed in the Czech national prostate cancer screening programme and discuss its initial outcomes, to provide insights for future screening.

## Patients and Methods

### Target population

The nationwide prostate cancer screening pilot targets men aged 50–69 years with no prior prostate cancer or clinical suspicion. Men over 70 are not actively enrolled but may be offered the same scheme based on individual health, at the physician’s discretion.

### Enrolment and diagnostic pathway

The screening algorithm was developed based on international evidence-based recommendations and the consensus of relevant professional societies [12–14], while incorporating local specifics of the Czech healthcare system. A detailed schematic representation of the algorithm is provided in Figure 1 and the Supplementary material.

**Figure 1.**
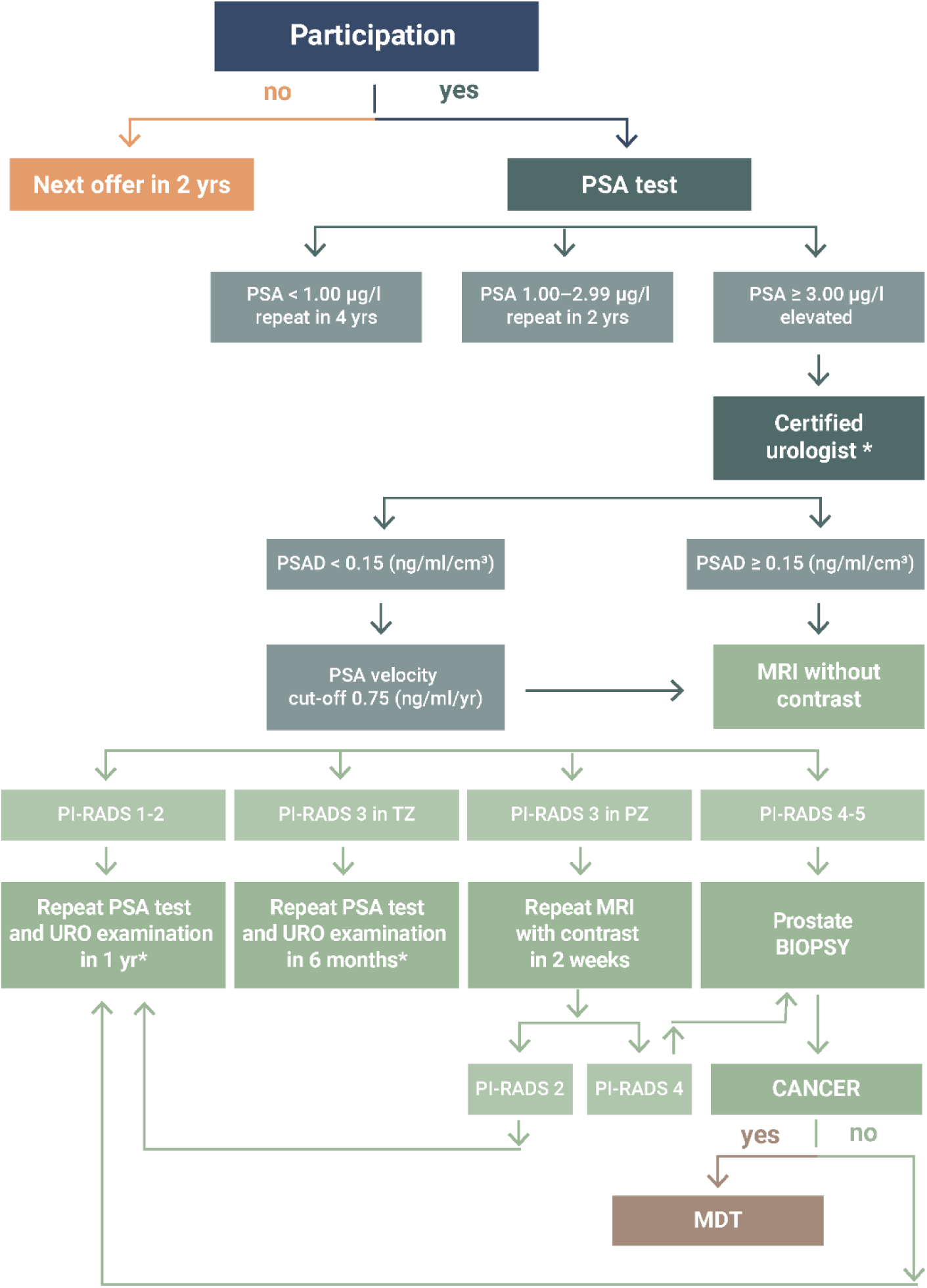
Screening algorithm: MRI = Magnetic Resonance Imaging; PI-RADS = Prostate Imaging Reporting and Data System; PSA = Prostate-Specific Antigen; PSAD = PSA Density; PZ = the peripheral zone; TZ = the transition zone; MDT = Multidisciplinary Team; *Includes a series of clinical examinations defined by the programme algorithm (Supplement).

Enrolment is undertaken by general practitioners (GPs) at routine preventive check-ups, recommended biennially in this age group. Urologists additionally enrol patients in the context of managing other urological conditions. After providing information and obtaining oral consent, a PSA test is sampled. Management is stratified by PSA: <1.0 µg/l, re-test in 4 years; 1.0–2.99 µg/l, in 2 years; ≥3.0 µg/l, referral to a Czech Urological Society–certified urologist for further assessment. If indicated (Fig. 1 and Supplement), men undergo biparametric MRI at a Ministry of Health–certified centre. MRI is interpreted using PI-RADS (Prostate Imaging Reporting and Data System): PI-RADS 1–2, repeat evaluation in 1 year; PI-RADS 3, repeat multiparametric MRI with contrast (peripheral zone lesion) or PSA re-test in 6 months (transitional zone lesion); PI-RADS 4–5, image fusion biopsy at a certified centre. Men with prostate cancer are referred to a multidisciplinary team, preferably at a certified onco-urological centre.

### Data sources and statistical methods

Patient-level data from the National Health Information System (NHIS) were used for monitoring and evaluation. The primary data source was the National Registry of Reimbursed Health Services (NRRHS), which collects data on reimbursed care from health insurance companies in the Czech Republic. Demographic data from the Czech Statistical Office (CZSO) were used to determine the target population.

The primary objective of the present analysis was the identification of men who were approached to enter the programme and their subsequent PSA values. As national registries do not currently allow the collection of quantitative values, it was only possible to obtain information on whether a man had a PSA value <1 µg/l, 1–2.99 µg/l, or ≥3 µg/l. All these results were also analysed by 5-year age groups. The involvement of GPs (in connection with preventive check-ups) and urologists was also monitored. The data on enrolment and baseline PSA testing were assessed for the first year of the organised programme, from January through December 2024. Men who were identified as having PSA ≥3 µg/l were analysed to see if they had a follow-up appointment with a urologist within 6 months. Due to data availability, this indicator was only assessed in men with a PSA result in January-June 2024.

The second objective was to analyse how the organised screening programme affects opportunistic PSA testing. Coverage of the target population was defined as the proportion of men aged 50–69 years with a PSA test in the past two years, divided by the number of men in the target population. Men who died before the year for which the indicator was evaluated were not included in the coverage. The age of men was determined for the year the coverage was evaluated, i.e., all men who turned 50 years of age in 2024 were categorised as aged 50 years.

The third objective was to assess diagnostic guidelines adherence before and after initiation of the screening programme. For men aged 50–69 years who underwent PSA testing (by GPs or urologists) in the first half of 2023 and the first half of 2024 (within or outside the screening programme), it was monitored whether they subsequently had a prostate biopsy and, if so, whether it was preceded by a prostate MRI, or the MRI was performed afterwards. Two-sided 95% confidence intervals (CI) were calculated for the presented proportions and their difference.

## Results

### Recruitment and baseline PSA testing

In the first twelve months of the Czech national pilot prostate cancer screening programme, a total of 150,498 men aged 50–69 years were offered to participate. Most men (97.1%) were enrolled in the screening programme by GPs, mainly at a routine preventive health check-up. In 2024, 3,291 GPs and 89 urologists enrolled men in the programme.

PSA results were available for 146,109 (diagram of participation flow in Supplementary material), whereas 1,501 men chose not to participate. For 2,888 men, information about their willingness to enter the programme was not available. Willingness to participate was calculated only for men for whom PSA values or non-participation was known (99.0%).

The largest 5-year age group was 50–54 years (33.5%). Over half of the men (56.5%, n = 82,613) had PSA <1.0 µg/l. The proportion of men with PSA <1.0 µg/l decreased with age, from 66.5% in men aged 50–54 to 44.4% in those aged 65–69 years. A PSA value ≥3.0 µg/l was identified in 8.8% (n = 12,874). This proportion increased with age, from 4.0% in the 50–54 age group to 15.7% in the 65–69 age group (Table 1).

**Table 1:** Age distribution of men approached for the screening programme and prostate-specific antigen (PSA) results by 5-year age groups. Data source: National Registry of Reimbursed Health Services, January-December 2024.

| Age (years) | 50–54 | 55–59 | 60–64 | 65–69 | Total |
| --- | --- | --- | --- | --- | --- |
| Screening offer, N (%) | 50,405<br>(33.5) | 38,103<br>(25.3) | 33,124<br>(22.0) | 28,866<br>(19.2) | 150,498<br>(100.0) |
| PSA screening results<br>available*, N | 48,988 | 37,033 | 32,176 | 27,912 | 146,109 |
| PSA <1.0 µg/l, N (%) | 32,599<br>(66.5) | 21,480<br>(58.0) | 16,137<br>(50.2) | 12,397<br>(44.4) | 82,613<br>(56.5) |
| PSA 1.0–2.9 µg/l, N (%) | 14,453<br>(29.5) | 12,798<br>(34.6) | 12,234<br>(38.0) | 11,137<br>(39.9) | 50,622<br>(34.6) |
| PSA ≥ 3 µg/l, N (%) | 1,936<br>(4.0) | 2,755<br>(7.4) | 3,805<br>(11.8) | 4,378<br>(15.7) | 12,874<br>(8.8) |

\*Screening results with known PSA values are presented. For less than 2% of men, the result is unknown due to incorrect reporting of data to healthcare payers, and another 1% chose not to be screened (Supplementary material: Figure 1).

Of the 6,595 men with a PSA result ≥3.0 µg/l between January and June 2024, 4,431 (67.2%) underwent a urological examination within six months of the PSA result. This proportion declined slightly with age, from 69.9% in the youngest 5-year age group to 64.8% in the oldest (Supplementary material: Table 1).

### Transition from opportunistic to organised screening: Results on a population level

During 2024, 53.3% of the target population of men aged 50–69 years without a prostate cancer diagnosis had a registered PSA test in the past 2 years, 5.9 percentage points more than in 2023 (47.4%; Figure 2).

**Figure 2.**
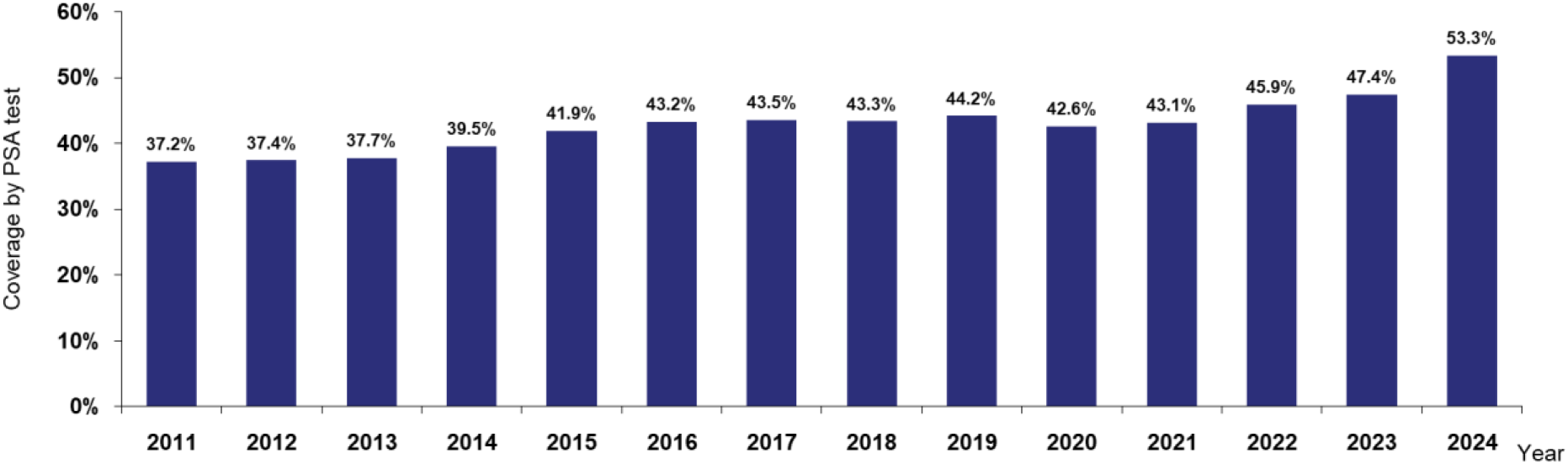
Coverage of the target population by the PSA test in a two-year interval in men aged 50–69 years. Data source: National Registry of Reimbursed Health Services; PSA = prostate-specific antigen

PSA testing coverage consistently increased across all age groups from 2023, the year before the organised screening programme, to its first year, 2024. The greatest increase was observed in the youngest age group (50–54 years), with coverage rising from 37.7% in 2023 to 44.6% in 2024 (Figure 3).

**Figure 3.**
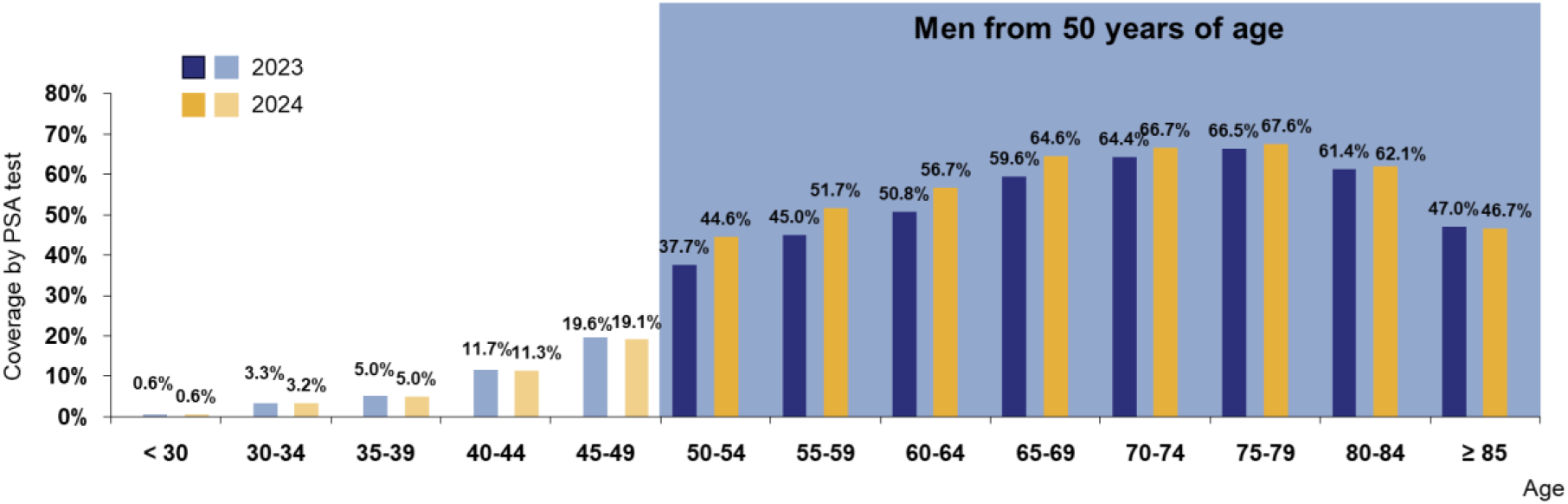
Target population coverage by age for a PSA test in the past two years, in 2023 and 2024. Data source: National Registry of Reimbursed Health Services; PSA = prostate-specific antigen

We assessed adherence to the screening diagnostic algorithm the first 6 months of the programme and compared with the same 6-month period before the implementation of the programme. The proportion of men undergoing prostate MRI before biopsy increased from 28.0% to 38.3%, a difference of 10.3 percentage points (95% CI 8.3–12.2 percentage points), while the rate of post-biopsy MRI decreased by 0.5 percentage points (Table 2).

**Table 2:** Comparison of the proportion of men with magnetic resonance imaging (MRI) before and after prostate biopsy, the first half of 2023 vs. the first half of 2024. Data source: National Registry of Reimbursed Health Services.

|  | January-June 2023 | January-June 2024 |
| --- | --- | --- |
| **Men with a PSA result, N** | 227,640 | 274,887 |
| PSA sampled in primary care, N | 119,107 | 164,606 |
| PSA sampled in urology, N | 108,533 | 110,281 |
| **Men with a prostate biopsy\*, N (%)** | 4,309 (1.9) | 4,670 (1.7) |
| PSA sampled in primary care, N (%) | 1,113 (0.9) | 1,504 (0.9) |
| PSA sampled in urology, N (%) | 3,196 (2.9) | 3,166 (2.9) |
| **Men with MRI before\*\* prostate biopsy, N (%)** | 1,208 (28.0) | 1,789 (38.3) |
| **Difference between periods, N (percent points; 95% CI)** | 10.3 (8.3–12.2) |  |
| PSA sampled in primary care, N (%) | 217 (19.5) | 464 (30.9) |
| PSA sampled in urology, N (%) | 991 (31.0) | 1,325 (41.9) |
| **Men with MRI after\*\*\* prostate biopsy, N (%)** | 360 (8.4) | 371 (7.9) |
| **Difference between periods, N (percent points; 95% CI)** | 0.5 (-1.5–0.7) |  |
| PSA sampled in primary care, N (%) | 147 (13.2) | 177 (11.8) |
| PSA sampled in urology, N (%) | 213 (6.7) | 194 (6.1) |

\* Biopsy taken within 180 days after PSA test.
\*\* MRI done between PSA test and prostate biopsy.
\*\*\* MRI done within 90 days after prostate biopsy.

## Discussion

This report presents the first results of an organised nationwide prostate cancer screening pilot programme in the Czech Republic, targeting men aged 50–69 years.

### Uptake of prostate cancer screening

Overall, 99.0% of the invited men decided to participate. This high participation rate is likely attributable to the fact that recruitment was conducted directly by GPs and urologists when men were physically attending healthcare and could have a PSA test with minimal extra effort. This personalized approach contrasts with many population-based screening programmes and trials that rely on centrally mailed invitation letters. It has been shown that men tend to have a higher level of trust in medical recommendations than in invitation letters [15].

Examples of population-based screening models that rely on mailed invitation letters are the Swedish Gothenburg-1 trial, where participation in the first rounds reached 59% among men aged 50–69 years [15], the Swedish regional organised prostate cancer testing (OPT) programmes with 35% participation among men aged 50 years [16], and the Finnish ProScreen trial with 51% participation among men aged 50–63 years [17].

The Czech approach builds on recommended regular preventive check-ups in primary care, enabling GPs to invite men for prostate screening during fully reimbursed visits, where PSA testing can be added with minimal extra burden. Urologist involvement in screening was limited due to payer contracts not automatically established by the end of 2024, delaying patient enrolment in the screening programme.

### PSA results

The observed proportion of 8.8% of men aged 50–69 years with PSA ≥3 µg/l was in the expected range, and so was the increase of this proportion with age [18]. In comparison, 6.7% of men aged 50–60 in the Gothenburg-2 trial [19], 12% of men aged 50–74 in the STHLM3-MRI trial [14], and 10.7% of men aged 50–74 in the Lithuanian population-based program had a PSA of ≥3 µg/l [20].

### Urology assessment

The proportion of men with PSA of ≥3 µg/l who undergo the recommended subsequent urological assessment was lower in the first six months of the Czech programme (67%) than in the four Swedish regions with a similar diagnostic pathway for OPT (83%–99%) [21]. The Czech proportion may be underestimated, as only a 6-month period was analysed and waiting times for appointments may sometimes be longer. One barrier to urological assessment is that it should be carried out by a certified urologist (lists are available on an official website [11]), who may be located some distance away from the participant’s residence. Another factor is that the Czech programme has no dedicated coordinators who actively communicate with participants and facilitate appointments. In contrast, Swedish OPT has regional offices that does this [21]. Consequently, continued participation in the Czech programme largely depends on individual motivation.

### Prostate MRI

The use of pre-biopsy MRI and targeted biopsy to reduce unnecessary systematic biopsy and overdiagnosis is crucial in modern prostate cancer screening [22]. After programme initiation in the Czech Republic, the overall proportion of men undergoing MRI before biopsy increased from 28.0% in the first half of 2023 to 38.3% in 2024, indicating improved alignment of diagnostic pathways.

MRI capacity is a known bottleneck when implementing screening [23]. In the Czech programme, an initial urology assessment is used to estimate risk and select men for MRI, in line with European Association of Urology recommendations and the PRAISE-U project [3] [13]. The impact of this pathway on MRI utilisation and diagnostic outcomes will be analysed once sufficient data are available. Standardised protocols are essential for ensuring high-quality MRI scanning and reading, and subsequent targeted biopsy sampling [23]. The Czech programme applies mandatory MRI protocols published by the Ministry of Health (Bulletin 15/2023).

### Change of coverage and practice

The observed increase in 2-year PSA testing coverage from 47.4% in 2023 to 53.3% in 2024 among men aged 50–69 years is likely an effect of the introduction of the organized screening programme. Although non-organized PSA testing still is practiced in the Czech Republic, the implementation of a systematic screening pathway has enhanced awareness and accessibility, especially through the engagement of GPs. The nearly 6 percentage point increase in coverage within a single year suggests successful early adoption of the programme.

In Lithuania, a primary care–based prostate cancer early detection programme was initiated in 2006 [20]. Sweden started implementation of regional, population-based organised testing (OPT) within public healthcare in 2020 [21]. This Swedish approach substantially increased the proportion of men aged 50 years undergoing PSA testing, from 14% to 35% [24]. Notably, unlike the Czech and Lithuanian programmes, the Swedish OPT relies on centralised invitations mailed to entire birth cohorts, without the involvement of general practitioners.

The planning of the Czech prostate cancer screening programme was facilitated by networking with the EU-funded PRAISE-U project [6]. PRAISE-U coordinates pilot projects in Poland, Ireland, Spain, Lithuania, and Estonia that are structured similarly to the Czech programme. The Italian Society of Urology has also proposed a structured model for PSA screening with defined roles for GPs and specialists [25]. Collaborative development of algorithms and shared performance standards are essential for effective prostate cancer screening in Europe.

### Strengths and limitations

The strengths of this report are the programme’s nationwide organised design, its full coverage by public health insurance, and its integration into routine care with enrolment at GPs’ preventive health check-ups. The programme focuses on clearly defined age groups, enhancing comparability and targeted evaluation.

Limitations include current lack of a national clinical register, restricting follow-up data such as biopsy results and urological assessments. Only 67% of men with elevated PSA were seen by a urologist within six months, which is suboptimal. A national register is under development. While initially observed uptake is promising, further efforts are needed to optimise informing the men, increase participation in follow-up steps, and assess its impact on reducing opportunistic PSA testing.

## Conclusions

In 2024, a nationwide prostate cancer screening programme was successfully launched in the Czech Republic. Early results show increased PSA testing coverage in men aged 50–69 years, particularly in the 50–54 age group, and greater use of pre-biopsy MRI in line with the diagnostic algorithm. These findings indicate that organised screening promotes both higher participation and more risk-adapted diagnostic practice. Recruitment through routine preventive check-ups in primary care, fully covered by public insurance, has proven feasible and establishes the foundation for future implementation of the routine programme. Further evaluation will further elaborate on the compliance of participants and providers with the recommended screening pathway.

## Data Availability

Aggregated data is available at Zenodo.

https://doi.org/10.5281/zenodo.18151231

## Data access

Hejcmanová, K. (2026). Czech nationwide prostate cancer screening pilot programme (DATA) [Data set]. Zenodo. https://doi.org/10.5281/zenodo.18151231

## Funding/Support and role of the sponsor

The study was supported by Programme JAC – project SALVAGE (CZ.02.01.01/00/22_008/0004644), financed by MEYS – Co-funded by the European Union.

The preparatory work for the nationwide pilot programme was supported by the project „Complex information background for improving the quality of cancer screening programmes within the National Screening Centre” (CZ.31.8.0/0.0/0.0/23_075/0008430) funded by the European Union from the Recovery and Resilience Facility through the National Recovery Plan of the Czech Republic.

## Acknowledgements

We would like to acknowledge close collaboration with the PRAISE-U project, which received funding from the EU4Health programme under grant agreement 101101217, co-funded by the European Union. Notably, we are grateful to Hendrik Van Poppel, Sarah Collen, and Monique Roobol-Bouts for the support of the Prague Prostaforum Declaration and preparatory works of the Czech programme. We also sincerely thank Eva Nevrtalová for her valuable consultations during the development of this manuscript and Markéta Vranová for her close collaboration in the joint coordination and implementation of the national programme in the Czech Republic.

## Supplementary information

Detailed description of the programme’s algorithm

Enrolment into the programme is facilitated through general practitioners (routine preventive health check-ups, which are recommended every two years for all individuals) and through urologists, who invite men already under follow-up for other urological conditions. In both settings, physicians provide education about importance of PSA testing, prostate cancer prevention and inform them about next steps of the programme. In accordance with Czech legislation (Act No. 372/2011), informed consent is needed but not necessarily in written form.

Information on participation is reported by physicians to health insurance companies and subsequently included in the National Registry of Reimbursed Health Services (NRRHS).

If the informed consent is given, a blood sample is collected to determine the serum PSA value.

Follow-up algorithm is based on PSA levels:

**PSA < 1.0 µg/l**: Repeat PSA screening in 4 years.

**PSA 1.0–2.99 µg/l**: Repeat PSA screening in 2 years.

**PSA ≥ 3.0 µg/l**: Considered a positive screening result – the man is referred for specialised urological evaluation by a certified urologist.

Certified urologist assessment (PSA ≥ 3.0 µg/l or suspicious digital rectal examination):

Certification is granted by the Czech Urological Society based on predefined criteria. In men with PSA ≥ 3.0 µg/l, a certified urologist carries out examinations according to the programme algorithm:

*repeat PSA test,*

*digital rectal examination,*

*ultrasonographic examination of the kidneys, urinary bladder,*

*transrectal ultrasound (TRUS) for prostate volume measurement,*

*calculation of PSA density (PSAD),*

*estimation of PSA velocity (PSAV) if more than one PSA test is available*.

Based on the findings (PSAD ≥ 0.15 or PSAV cut-off 0.75 ng/ml/year), the urologist may refer for magnetic resonance imaging (MRI), either biparametric (bpMRI, contrast-free) or multiparametric (mpMRI, with intravenous contrast), at an MRI unit certified by the Ministry of Health.

In the case of PSA >1 µg/l and a suspicious digital rectal examination, the urologist can initiate either an MRI or a prostate biopsy.

MRI Evaluation within the programme (PI-RADS — the Prostate Imaging Reporting and Data System):

**PI-RADS 1 and 2 (negative)**: Further management follows the approved algorithm. The man is returned to certified urologist for, urological examination with a repeat MRI scheduled in one year.

**PI-RADS 3 (equivocal)** in the peripheral zone (PZ): Repeat MRI with intravenous gadolinium-based contrast enhancement (mpMRI) within two weeks. If then PI-**RADS 2 findings**, repeat PSA testing and urological examination after one year. If **PI-RADS 4**, targeted biopsy with MRI/ultrasound image fusion.

**PI-RADS 3** (equivocal) in the transitional zone (TZ): Repeat PSA test and urological examinations (TRUS, PSAD, PSAV) after six months; additional mpMRI or prostate biopsy (high suspicion) if considered indicated.

**PI-RADS 4 and 5 (positive)**: MRI/ultrasound fusion biopsy at a certified biopsy centre. Certification is granted by the Ministry of Health based on established criteria.

Men diagnosed with prostate cancer are referred to a multidisciplinary team for further management. If the biopsy is negative, follow-up MRI is scheduled after one year.

**Supplementary Table 1:** Proportion of men with PSA ≥ 3 µg/l and a follow-up examination by a urologist within 6 months by 5-year age group. Data source: National Registry of Reimbursed Health Services January-June 2024.

| Age (years) | 50–54 | 55–59 | 60–64 | 65–69 | Total |
| --- | --- | --- | --- | --- | --- |
| Men with PSA $\geq 3$ $\mu\text{g/l}$, N | 1,000 | 1,412 | 1,945 | 2,238 | 6,595 |
| Men with PSA $\geq 3$ $\mu\text{g/l}$ examined by urologist, N (%) | 699<br>(69.9) | 964<br>(68.3) | 1,317<br>(67.7) | 1,451<br>(64.8) | 4,431<br>(67.2) |

**Supplementary Figure 1:**
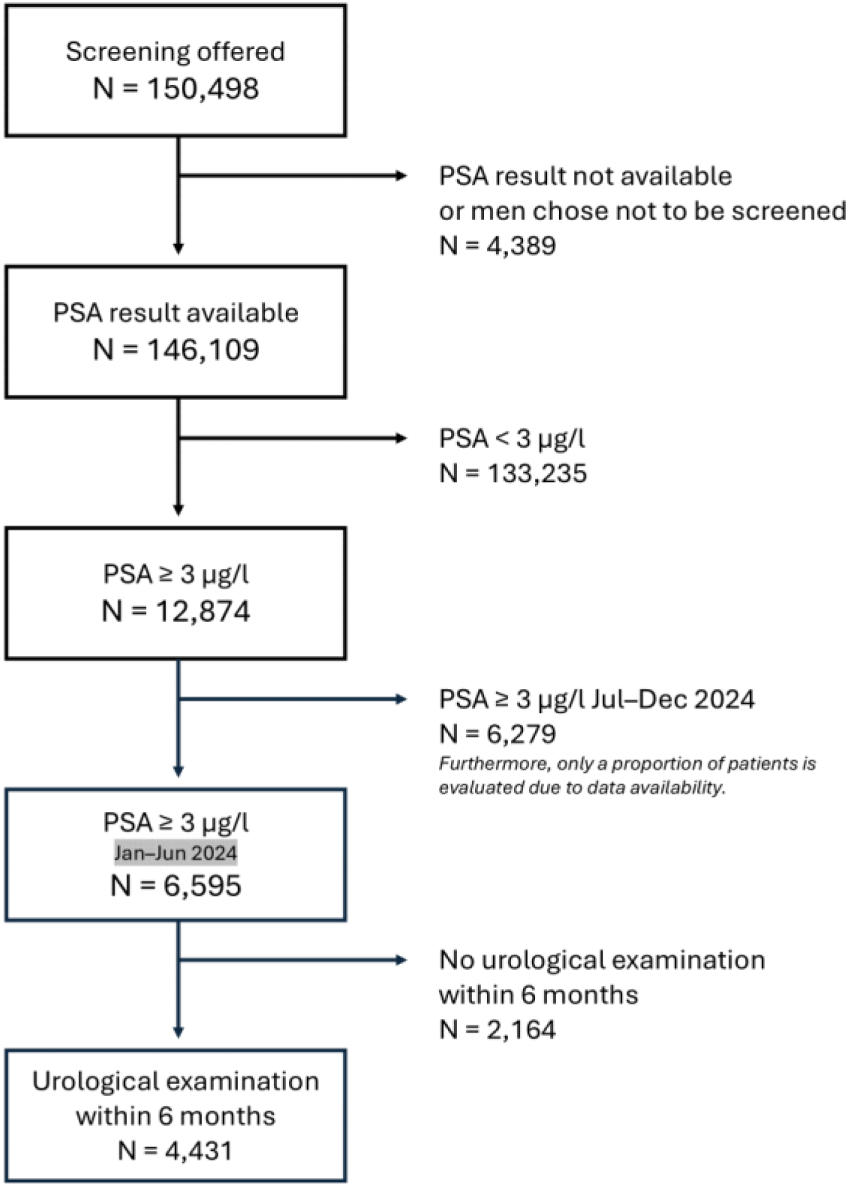
Participation rate, PSA results in first 12 months, and follow-up examination by a urologist within 6 months. Data source: National Registry of Reimbursed Health Services January-June 2024.

## Take-home Message

Organised prostate cancer screening is feasible at the national level. Early results show increasing population coverage and better alignment with evidence-based diagnostic practices. Active involvement of GPs and urologists is crucial for improving screening uptake and adherence to guidelines.

## Notes

### Competing Interest Statement

The authors have declared no competing interest.

### Funding Statement

The study was supported by Programme JAC project SALVAGE, financed by MEYS Co–funded by the European Union. The preparatory work for the nationwide pilot programme was supported by the project Complex information background for improving the quality of cancer screening programmes within the National Screening Centre funded by the European Union from the Recovery and Resilience Facility through the National Recovery Plan of the Czech Republic.

### Author Declarations

Only aggregated and fully anonymized data were used for the purposes of the publication. At no point of research did the authors have access to individual-level, non-anonymized participant data. All data handling activities are carried out under the supervision of the Data Handling Committees of the Institute of Health Information and Statistics (ÚZIS), which include a designated Data Protection Officer responsible for oversight of work with sensitive data.

### Summary of Updates

The SALVAGE project reqiures ORCID numbers, which were joined.

## References

[1] Sung H, Ferlay J, Siegel RL, Laversanne M, Soerjomataram I, Jemal A, et al. Global Cancer Statistics 2020: GLOBOCAN Estimates of Incidence and Mortality Worldwide for 36 Cancers in 185 Countries. CA Cancer J Clin 2021;71:209–49. 10.3322/caac.21660.

[2] Krejčí D, Mužík J, Šnábl I, Gregor J, Komenda M, Dušek L. Portal of Cancer Epidemiology in the Czech Republic [online] n.d. https://www.svod.cz.

[3] Van Poppel H, Hogenhout R, Albers P, van den Bergh RCN, Barentsz JO, Roobol MJ. Early Detection of Prostate Cancer in 2020 and Beyond: Facts and Recommendations for the European Union and the European Commission. Eur Urol 2021;79:327–9. 10.1016/j.eururo.2020.12.010.

[4] Roobol MJ, de Vos II, Månsson M, Godtman RA, Talala KM, den Hond E, et al. European Study of Prostate Cancer Screening - 23-Year Follow-up. N Engl J Med 2025;393:1669–80. 10.1056/NEJMoa2503223.

[5] Arnsrud Godtman R, Holmberg E, Lilja H, Stranne J, Hugosson J. Opportunistic Testing Versus Organized Prostate-specific Antigen Screening: Outcome After 18 Years in the Göteborg Randomized Population-based Prostate Cancer Screening Trial. Eur Urol 2015;68:354–60. 10.1016/j.eururo.2014.12.006.

[6] Chandran A, van Harten M, Singh D, Vilaseca J, Patasius A, Tupikowski K, et al. Risk-stratified Approach to Implementing Population-based Prostate Cancer Screening in Five Pilot Sites in the European Union: A Protocol for the PRAISE-U Project. Eur Urol Open Sci 2024;70:8–17. 10.1016/j.euros.2024.09.003.

[7] Vynckier P, Annemans L, Raes S, Amrouch C, Lindgren P, Májek O, et al. Systematic Review on the Cost Effectiveness of Prostate Cancer Screening in Europe. Eur Urol 2024;86:400–8. 10.1016/j.eururo.2024.04.036.

[8] Beyer K, Leenen R, Venderbos LDF, Helleman J, Denijs F, Bramer W, et al. Health Policy for Prostate Cancer Early Detection in the European Union and the Impact of Opportunistic Screening: PRAISE-U Consortium. J Pers Med 2024;14:84. 10.3390/jpm14010084.

[9] Májek O, Babjuk M, Roobol MJ, Bratt O, Van Poppel H, Zachoval R, et al. How to follow the new EU Council recommendation and improve prostate cancer early detection: the Prostaforum 2022 declaration. Eur Urol Open Sci 2023;53:106–8. 10.1016/j.euros.2023.05.011.

[10] Leenen RCA, Venderbos LDF, Helleman J, Gómez Rivas J, Vynckier P, Annemans L, et al. Prostate Cancer Early Detection in the European Union and UK. Eur Urol 2025;87:326–39. 10.1016/j.eururo.2024.07.019.

[11] Koudelková M, Zachoval R, Ferda J, Babjuk M, Matoušková M, Herber O, et al. Czech nationwide prostate cancer screening pilot programme. Prostascreening.cz 2024. https://www.prostascreening.cz/ (accessed August 6, 2025).

[12] Kohestani K, Chilov M, Carlsson SV. Prostate cancer screening-when to start and how to screen? Transl Androl Urol 2018;7:34–45. 10.21037/tau.2017.12.25.

[13] Van Poppel H, Hogenhout R, Albers P, van den Bergh RCN, Barentsz JO, Roobol MJ. A European Model for an Organised Risk-stratified Early Detection Programme for Prostate Cancer. Eur Urol Oncol 2021;4:731–9. 10.1016/j.euo.2021.06.006.

[14] Eklund M, Jäderling F, Discacciati A, Bergman M, Annerstedt M, Aly M, et al. MRI-Targeted or Standard Biopsy in Prostate Cancer Screening. N Engl J Med 2021;385:908–20. 10.1056/nejmoa2100852.

[15] Svensson L, Bratt O, Hugosson J, Stinesen K. Prostate Cancer Screening Decisions: Which Aspects Do Men Value Most? An Interview Study With Men Invited to a Population-Based Program. Am J Mens Health 2025;19:15579883251344563. 10.1177/15579883251344563.

[16] Bratt O, Godtman RA, Jiborn T, Wallström J, Akre O, Carlsson S, et al. Population-based Organised Prostate Cancer Testing: Results from the First Invitation of 50-year-old Men. Eur Urol 2024;85:207–14. 10.1016/j.eururo.2023.11.013.

[17] Auvinen A, Tammela TLJ, Mirtti T, Lilja H, Tolonen T, Kenttämies A, et al. Prostate Cancer Screening With PSA, Kallikrein Panel, and MRI: The ProScreen Randomized Trial. JAMA 2024;331:1452–9. 10.1001/jama.2024.3841.

[18] Deantoni EP. Age-Specific Reference Ranges for PSA in the Detection of Prostate Cancer. Oncology 1997;11:475–85.

[19] Hugosson J, Månsson M, Wallström J, Axcrona U, Carlsson SV, Egevad L, et al. Prostate Cancer Screening with PSA and MRI Followed by Targeted Biopsy Only. N Engl J Med 2022;387:2126–37. 10.1056/nejmoa2209454.

[20] Patasius A, Krilaviciute A, Smailyte G. Prostate cancer screening with psa: Ten years’ experience of population based early prostate cancer detection programme in Lithuania. J Clin Med 2020;9:1–9. 10.3390/jcm9123826.

[21] Bratt O, Butt ST, Carlsson C, Jelf-Eneqvist L, Gunnarsson O, Ihre A, et al. Swedish regional population-based organised prostate cancer testing: why, what and how? Scand J Urol 2025;60:97–104. 10.2340/sju.v60.43809.

[22] Schoots IG, Ahmed HU, Albers P, Asbach P, van den Bergh RCN, Godtman RA, et al. Magnetic Resonance Imaging–based Biopsy Strategies in Prostate Cancer Screening: A Systematic Review. Eur Urol 2025;88:247–60. 10.1016/j.eururo.2025.05.038.

[23] Singh D, Chandran A, Panebianco V, Vilaseca J, Patasius A, Tupikowski K, et al. MRI capacity assessment for prostate cancer screening in five sites of Europe. Eur J Radiol 2025;190:112235. 10.1016/j.ejrad.2025.112235.

[24] Järbur E, Holmberg E, Björk-Eriksson T, Bratt O, Arnsrud Godtman R. Associations between socioeconomic factors and PSA testing in a population-based organised testing programme and routine healthcare: a register-based study of 50-year-old men. BMJ Oncol 2024;3:e000400. 10.1136/bmjonc-2024-000400.

[25] Ficarra V, Italian Society of Urology (SIU) panel, Bartoletti R, Borghesi M, Caffo O, De Nunzio C, et al. Organized prostate cancer screening program: a proposal from the Italian Society of Urology (SIU). Minerva Urol Nephrol 2024;76. 10.23736/s2724-6051.24.06117-2.

